# SYSTEMATIC REVIEW ON EPIDEMIOLOGY, INTERVENTIONS AND MANAGEMENT OF NONCOMMUNICABLE DISEASES IN ACUTE AND EMERGENCY CARE SETTINGS IN KENYA

**DOI:** 10.1101/2020.06.25.20138396

**Authors:** Christine Ngaruiya, Annrita Kawira, Florence Mali, Faith Kambua, Beatrice Mwangi, Mbatha Wambua, Denise Hersey, Laventa Obare, Rebecca Leff, Benjamin Wachira

## Abstract

**Introduction:** Mortality and morbidity from Non-Communicable Diseases (NCDs) in Africa are expected to worsen if the status quo is maintained. Emergency care settings act as a primary point of entry into the health system for a spectrum of NCD-related illnesses, however, there is a dearth of literature on this population. We conducted a systematic review assessing available evidence on epidemiology, interventions and management of NCDs in acute and emergency care settings in Kenya, the largest economy in East Africa and a medical hub for the continent.

**Methods:** All searches were run on July 15, 2015 capturing concepts of NCDs, and acute and emergency care. The study is registered at PROSPERO (CRD42018088621).

**Results:** We retrieved a total of 461 references, and an additional 23 articles in grey literature. 391 studies were excluded by title or abstract, and 93 articles read in full. We included 10 articles in final thematic analysis. The majority of studies were conducted in tertiary referral or private/mission hospitals. Cancer, diabetes, cardiovascular disease and renal disease were addressed. Majority of the studies were retrospective, cross-sectional in design; no interventions or clinical trials were identified. There was a lack of access to basic diagnostic tools, and management of NCDs and their complications was limited.

**Conclusion:** There is a paucity of literature on NCDs in Kenyan emergency care settings, with particular gaps on interventions and management. Opportunities include nationally representative, longitudinal research such as surveillance and registries, as well as clinical trials and implementation science to advance evidence-based, context-specific care.

Key questions

What is already known?

- Mortality and morbidity from Non-Communicable Diseases (NCDs) in Africa are expected to worsen if the status quo is maintained.
- Emergency care settings act as a primary point of entry into the health system for a spectrum of NCD-related illnesses. However, the ED as a primary source of NCD care has been under-prioritized in LMICs.

What are the new findings?

- There is limited evidence on NCDs in Kenyan emergency care settings, with particular gaps in interventions and management.
- The majority of studies were conducted in tertiary referral or private/mission hospitals in urban settings, which are not accessed by the majority of Kenyans residing in rural areas.
- Access to basic diagnostic tools, and management of NCDs and their complications is limited for Kenyans in emergency care settings.

What do the new findings imply?

- To guide intervention development and improve health outcomes for NCDs in Kenya, Kenya will need to prioritize primary data collection in the ED setting with special attention to emergency care at lower acuity health facilities and in rural areas.
- More rigorous research and reporting on effective strategies for delivering NCD care in acute and emergency care populations is urgently needed, given the increasing burden of NCDs in LMICs, including in Kenya.

## INTRODUCTION

Non-Communicable Diseases (NCDs) contribute to a global threat accounting for nearly 75% of deaths globally (1). Furthermore, mortality and morbidity from NCDs in Africa, as measured by Disability Adjusted Life Years (DALYs), has increased by 67% in recent decades and this burden is only expected to increase (2, 3). The leading causes of mortality due to NCDs are: cardiovascular disease, cancer, diabetes, and chronic respiratory disease such as asthma and Chronic Obstructive Pulmonary Disease (COPD)(4). Addressing the NCD epidemic will require efforts across the healthcare system. The Emergency Department (ED) acts as a primary access point for entry into the system (5, 6). Patients present with a variety of NCD-related complaints, and are from across the age spectrum (7). It is estimated that 24 million lives and nearly 1 billion DALYs could be averted by advancement of emergency care in LMICs (8). By targeting acute and emergency care settings, we can affect disease in a large proportion of the population that may not otherwise access care. In order to address NCDs in acute and emergency care settings in LMICs, primary data is crucial (7-11). Our team conducted a systematic review assessing available evidence on epidemiology, interventions and management of NCDs in acute and emergency care settings in Kenya. Through this study, we aim to inform research priorities, clinical intervention development and local policy implementation. This is the first study of its kind in the region providing a comprehensive assessment on literature addressing NCDs in emergency care populations.

## METHODS

### Protocol and registration

The protocol of the present a systematic review is registered under PROSPERO (CRD42018088621)(9).

### Inclusion criteria

We included studies that constituted epidemiology, interventions and management that addressed acute or emergency care populations. Given patients from the ED are often triaged to the outpatient clinics, even if they present to the ED, in order to be comprehensive we have included terms that account for both settings that capture potential acute patient populations.

### Exclusion criteria

Studies that did not involve a Kenyan population or occur in Kenya, that did not involve an acute or emergency care/ emergency department population, or that did not constitute an epidemiological study, intervention or management pertinent to acute or emergency care populations were excluded. No exclusions or restrictions were applied relative to publication date and language.

### Search Strategy and Study selection

The literature search was completed by an expert medical librarian (DH) and lead author (CN), with input from senior author (BW) for context-specific terminology. All searches were run on July 15, 2015. The databases searched were MEDLINE (OvidSP 1946-July Week 1 2015), MEDLINE (PubMed, for in-process and non-indexed citations), Embase (OvidSP 1974-2015 July 14), Scopus (Elsevier, all years), Web of Science Core Collection (Thomson Reuters, 1964-present), Africa-Wide Information (EBSCOhost, all years), and CINAHL (EBSCOhost, all years). In addition, a search of grey literature was performed in both the Intergovernmental Organization and Nongovernmental Organization Google Custom Search Engines and Greylit.org. The formal search strategies used relevant controlled vocabulary terms and synonymous free text words and phrases to capture concepts of non-communicable diseases “heart attack”, “myocardial ischemia”, “acute coronary syndrome”, “hypertension”, “COPD”, “diabetes”, “asthma”), and Emergency Medicine (“acute care”, “emergency services”, “emergency department”). For all items identified, the titles and abstracts were independently screened by two reviewers (AK, FM, BM, or FK.). For all items deemed potentially eligible, screening of full texts was performed by the same reviewers.

### Data Extraction

Two independent reviewers performed data extraction. Disagreement was solved by direct discussion guided by the lead author CN. Final synthesis, as described in our study registration (9), was guided by the outcomes: (1) breakdown by study type i.e. epidemiology, intervention, management, (2) outcomes from individual level study on effects of intervention, where applicable, (3) breakdown by disease focus of study (Cardiovascular disease, chronic respiratory disease, diabetes, cancer, and other), and (4) other pertinent emergent themes from individual studies.

## RESULTS

The final searches retrieved a total of 510 references, which were pooled in Endnote and de-duplicated to 461 total articles (see Fig 1). 391 studies were excluded by title or abstract and 93 articles were read in full to further screen for inclusion in final thematic analysis. An additional 217 articles were identified in grey literature, and 23 articles were included in the final review. Out of the studies screened, ten were included in the final analysis. Primary reasons for exclusion were that studies were conducted outside of Kenya or did not include Kenyan populations (such as publications conducted in other African populations that were published in Kenyan academic journals).

**Fig 1.**
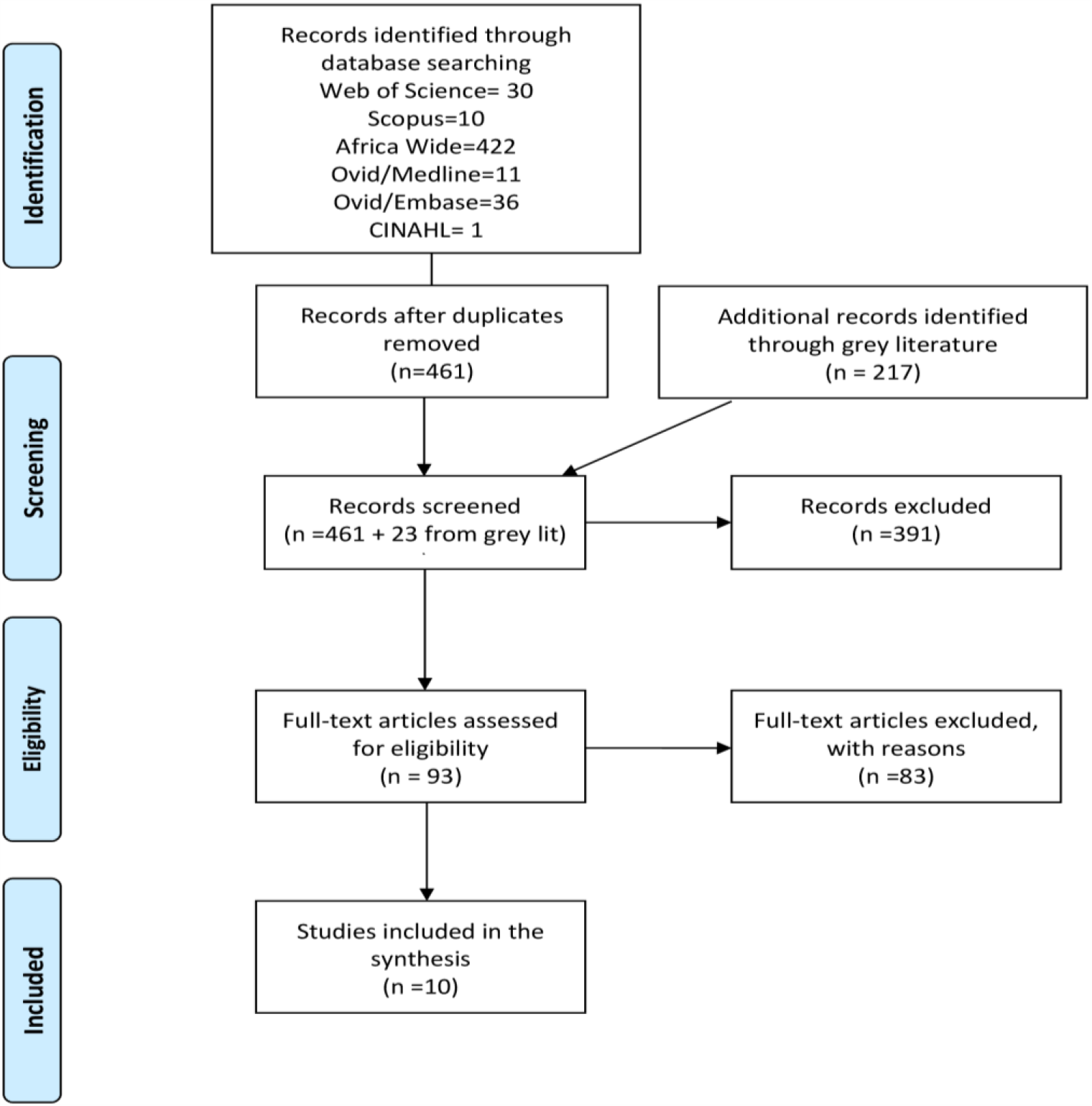
PRISMA Diagram. From: Moher D, Liberati A, Tetzlaff J, et al. The PRISMA Group. Preferred Reporting Items for Systematic Reviews and Meta-Analyses: The PRISMA Statement. *PLoS Med* 2009;6: e1000097. doi:10.1371/journal.pmed1000097

Additionally, several references were policy documents or concept papers with no primary data collection. Systematic reviews and meta-analyses were also excluded from our final analysis. Finally, studies that were basic science in nature, or that lacked clinical relevance in acute or emergency care of patients were excluded (such as use of breast cancer markers for guiding long-term management of treatment from an outpatient cancer clinic sample population). Studies that addressed trauma or traumatic injuries were also excluded given the review scope being specific to chronic NCDs. All ten articles were summarized by key concepts and themes (see Table 1).

**Table 1.** Results of a Systematic Review on Epidemiology, Interventions and Management of Noncommunicable Diseases in Acute and Emergency Care Settings in Kenya

| Author name and Year | Population | Study type | Dates | Sample size (n) | Study primary findings |
| --- | --- | --- | --- | --- | --- |
| **Burke et al (2014)** | All 30 primary and secondary hospitals and a stratified random sampling of 30 dispensaries and health centres in western Kenya | Cross-sectional | Nov 2013-Jan 2014 | 60 (healthcare facilities) | Top ten most common conditions were reported to be communicable diseases, except for trauma (30%-60%, depending on facility). 100% of MI patients were referred in lower-level facilities, 80% in higher-level facilities. 93% felt ill-prepared to manage DKA at lower-level facilities, 67% at higher-level facilities. Among diagnostics: 87% had BP cuffs, 13% had EKG, 30% had XR, 57% had glucometer. Among medications and management supplies: 12% had nitroglycerin, 48% had insulin, 53% had oxygen, 38% had intubation supplies. |
| **Dawsey et al (2010)** | Patients with a histologic or endoscopic diagnosis of EC who were <30y | Case series | 1996-2009 | 109 (65 male; 44 female) | Follow-up information obtained on n=60 (55%) of patients. 4 (7%) were candidates for esophagectomy, one of whom survived >5 years. N=45 (79%) had positive family history of CA, n=21 (43%) had family history of esophageal CA. Tobacco smoking present in n=9 (15%), alcohol use in n=9 (15%). |
| **Nkumbe et al (2010)** | Diabetes clinic patients, diagnosed with DM in past 12 months | Cross-sectional | 2001-2002 | 71 (30 male; 41 female) | A total of n=65/71 patients were included, with the remainder excluded due to poor quality of images obtained. 42% of subjects were male. The prevalence of diabetic retinopathy in men and women was 33% and 28% respectively ($P = 0.15$), while the overall prevalence was 30.4%. Diabetic retinopathy was unilateral in three patients (6.5%) and bilateral in 11 (23.9%). Four out of 46 patients (8.7%) had clinically significant macula edema (CSME). |
| **Aduda et al (2014)** | Women > 18y who had been tested for Syphilis or undergone cervical cancer screening | Qualitative | 2009 | 64 (females) | On cervical cancer screening, there was understanding of asymptomatic nature of disease in early stages, and on typical signs and symptoms later, but also misconceptions on how it is contracted. Respondents were appreciative of pre-test consent and found it educational, but still found screening fearful with concerns about possible positive screening results. Results were desired as soon as possible, and verbal administration of results was preferred over written given anxiety, impersonality, and illiteracy. A deference to "God's will" was also evident. |
| **Otieno et al (2010)** | Emergency Department and hospital patients | Cross-sectional | Dec 2001-Aug 2002 | 51 (25 male; 22 female) | 8% (n=51/648) of diabetic patients admitted during study period had DKA. Mean age 33.4 (95% CI 15.2). 51% were newly diagnosed. Polyuria (85%), polydipsia (83%) and vomiting (43%) were lead presenting symptoms. 36% (n=17) were obtunded with GCS 3-8. 32% (n=15) were severely dehydrated. Mortality was 30% (n=14/47). Abnormal renal function (71%), AMS (100%), new DM diagnosis (64%) and being female (64%) were poor prognosticators. |
| **Wachira et al (2012)** | Emergency Department patients presenting alive, on randomly generated set of days during data | Cross-sectional | Oct 2010-Dec 2010 | 1887 (940 male; 947 female) | 27% of patients aged 0-9y; 70% aged >13y. 58% had tests done, malaria blood film most common. Trauma lead diagnoses in adults (24%), malaria in children (24%). Wound care (26%), fluid resuscitation (10%), management of bronchospasm (7%), splinting (4%), and management of hyperglycemia (3%) were top 5 most common treatments. 19% (n=354) were admitted, 7% (n=127) were referred. |

|  | collection period |  |  |  |  |
| --- | --- | --- | --- | --- | --- |
| **Wachira et al (2014)** | Adults >21y admitted from the ED with a diagnosis of Acute Coronary Syndrome (ACS) | Retrospective chart review | Jan 2012-Feb 2013 | 45 (37 male; 8 female) | Compliance rates with: ECG done in 10 min of first medical contact was 89%; door-to-needle (fibrinolysis) goal of 30 min was 43%; door-to-balloon (PCI) goal 90 min was 29%; in-hospital complication rate was 13.3%. |
| **Wachira et al (2015)** | Adults aged >18y with In-Hospital Cardiac Arrest (IHCA) | Retrospective chart review | Jan 2013-Dec 2013 | 108 (63 male; 45 female) | The predominant initial rhythms post cardiac arrest were pulseless electrical activity (41.7%) or asystole (35.2%). Hypertension (43.5%), septicemia (40.7%), renal insufficiency (30.6%), diabetes mellitus (25.9%) and pneumonia (15.7%) were the leading pre-existing conditions in the patients. A Modified Early Warning Score (MEWS) of 5 or more was reached in 56 (67.5%, n = 83) patients before the cardiac arrest. The rate of survival to hospital discharge was 11.1%. All the patients who survived to hospital discharge had a good neurological outcome. |
| **White et al (2002)** | All cancers diagnosed during the study period at Tenwek Hospital | Case series | Jan 1989-Dec 1998 | 1459 | Top 5 most commonly diagnosed cancers: esophageal (n=274), stomach (n=137), prostate (n=79), liver (n=71) and colorectal (n=61). Breast cancer among top ten (n=51). Mean age at diagnosis was: 56y for women and 54y for men (14-91y). 11% of esophageal cancer cases were aged <30y. |
| **Wools-Kaloustian et al (2007)** | Outpatient HIV clinic patients | Cross-sectional | May 2004-Nov 2005 | 389 (125 male; 264 female) | For the 373 individuals with complete data, the mean CrCl was 90ml/min (range 30 –200ml/min). CrCl <60ml/min was identified in 43 (11.5%) subjects with 18 (4.8%) having a CrCl <50ml/min. In the multi-variate analysis, only lower haemoglobin [OR 0.79 (0.69–0.91), p=0.001] and wasting syndrome [OR 4.17 (1.15–15.11), p=0.03] were associated with a CrCl <60ml/min. History of tuberculosis [OR 3.0 (1.02–8.96), p=0.04] was significantly associated with proteinuria. |

Abbreviations: ACS-Acute Coronary Syndrome, AMS-Altered Mental Status, BP-Blood Pressure, CA-Cancer, DM-Diabetes Mellitus, EC-Esophageal Cancer, ED-Emergency Department, EKG- Electrocardiogram, DKA-Diabetic Ketoacidosis, GCS-Glasgow Coma Scale, IHCA-In-Hospital Cardiac Arrest, NCD-Non-Communicable Disease, XR- X-ray

### Results by study design and disease focus

Among all of the studies included for final review (n=10), majority were quantitative in study design, and only one was qualitative in nature. Burke et al assessed capacity for NCD care in a sample of facilities in the Western region of Kenya (10). Wachira et al conducted a descriptive analysis on all patients accessing care in a sample of 15 EDs (11). Two studies assessed cardiovascular disease patient outcomes at a private, tertiary level hospital (12, 13).

Otieno et al assessed outcomes of Diabetic Ketoacidosis (DKA) patients admitted at the national public, tertiary referral hospital (14). Nkumbe et al assessed prevalence of diabetic retinopathy at an outpatient ophthalmology clinic (15). Two case series were conducted at a mission hospital: one included all cancer patients and a second focused solely on esophageal cancer patients (16, 17). Wools-Kaloustian conducted a cross-sectional study at an HIV clinic in Western Kenya to assess prevalence of renal disease in these patients (18).

Finally, a qualitative study was conducted by Aduda et al in an outpatient clinic to assess ethical considerations of screening for cervical cancer among patients (19). Of the ten studies, those with a specific disease focus were: cardiovascular disease (n=2), cancer (n=3), diabetes (n=2) and renal disease (n=1). An assessment of the risk of bias of the included studies are included(see Table 2).

**Table 2.** Quality Assessment (Method: Cochrane Collaboration’s Tool for Assessing Risk of Bias)

|  | Random sequence generation (selection bias) | Allocation concealment (selection bias) | Blinding of participants and personnel (performance bias) | Blinding of outcome assessment (detection bias) | Incomplete outcome data addressed (attrition bias) | Selective reporting (reporting bias) | Other sources of bias | Total<br>Low on Risk of Bias |
| --- | --- | --- | --- | --- | --- | --- | --- | --- |
| **Burke et al (2014)** | Low risk | Low risk | High risk | High risk | Low risk | Low risk | Low risk | (5/7) |
| **Dawsey et al (2010)** | Low risk | Low risk | Low risk | Low risk | High risk | High Risk | Low Risk | (5/7) |
| **Nkumbe et al (2010)** | Low risk | Low risk | Low risk | Low risk | High risk | Low risk | Low risk | (6/7) |
| **Aduda et al (2014)** | High risk | High risk | High risk | Low risk | High risk | Low risk | Low risk | (3/7) |
| **Otieno et al (2010)** | High risk | Low risk | High risk | Low risk | Low risk | Low risk | Low risk | (5/7) |
| **Wachira et al (2012)** | Low risk | Low risk | Low risk | Low risk | Low risk | Low risk | Low risk | (7/7) |
| **Wachira et al (2014)** | Low risk | Low risk | Low risk | Low risk | Low risk | Low risk | Low risk | (7/7) |
| **Wachira et al (2015)** | Low risk | Low risk | Low risk | Low risk | High risk | Low risk | Low risk | (6/7) |
| **White et al (2002)** | Low risk | Low risk | Low risk | Low risk | High risk | Low risk | Low risk | (6/7) |
| **Wools-Kaloustian et al (2007)** | High risk | High risk | Low risk | Low risk | Low risk | Low risk | Low risk | (5/7) |

From: Higgins JPT, Altman DG, Sterne JAC, eds. Chapter 8: Assessing risk of bias in included studies. In: Higgins JPT, Green S, eds. Cochrane Handbook for Systematic Reviews of Interventions. Version 5.1.0 [updated March 2011]. The Cochrane Collaboration, 2011.

### Results by target population focus and population location

Only five studies addressed ED patients or patients admitted to the hospital from the ED. Three of the ED-specific studies incorporated patient populations in the public health system (10, 11, 14), where the majority of patients in Kenya receive care. The other two studies on ED populations (12, 13) were among patients at a private teaching and referral hospital, Aga Khan University Hospital – Nairobi (AKUHN).

Of the five additional studies that were not conducted on ED patients, two were conducted in the outpatient setting at a well-established mission hospital, Tenwek hospital (16, 17). The final three were conducted at specialty clinics: an outpatient diabetes clinic(15), an outpatient HIV clinic(18) and among cervical cancer patients at an antenatal clinic (19).

Of note, none of the studies were nationally representative in sampling. Women were equitably represented in the studies or constituted the predominant sample population. Results on children were only present in one of the studies (11). Finally, half of the studies were conducted at facilities in the urban capital city, Nairobi.

### Results by individual study

In the capacity assessment by Burke et al of 60 healthcare facilities in Western Kenya, their capacity to manage NCDs or complications of NCDs (heart attack and diabetic ketoacidosis) was poor (10). Majority of patients required referral for heart attack, and even at higher acuity level facilities in the sample, only 44% reported stabilizing heart attack patients prior to transfer, such as providing oxygen. In a national sample of 15 public hospitals in Kenya by Wachira et al, they described patient presentations and management (11). Hypertension (3%) and acute asthma attack (3%) were among the top ten lead diagnoses in adults. Among the top most common treatments were “management of acute bronchospasm” (7%) and “management of hyperglycemia” (3%). Malaria screening was the most common test ordered.

A retrospective chart review was also conducted by Wachira et al among ED patients accessing care at the private hospital AKUHN with the aim of assessing compliance with guidelines for the management of ST-elevation Myocardial Infarction (STEMI)(12). They found the prevalence of compliance ranged from 29%-89%, with the goal of initial EKG being done within 10 minutes of first medical contact being the most successful intervention (see Table 1). Compliance with guidelines was lower for definitive treatment, with “door-to-needle” (fibrinolysis) goal achieved only 43% of the time, and “door-to-balloon” primary percutaneous coronary intervention (PCI) goal achieved only 29% of the time. There was near universal administration of aspirin (98%) and clopidogrel (91%), though time to administration was not addressed.

The second study on cardiovascular disease was a descriptive analysis of In-Hospital Cardiac Arrest (IHCA) patients, which was also a retrospective chart review conducted at AKUHN (13). The review included all patients seen in 2013. 108 patients were included in the study. Only 4% of arrests occurred in the ED, and the majority (40%) occurred in the Intensive Care Unit (ICU). In case of an IHCA, an assigned team of ED and ICU nurses and doctors responds. The average team response time was 164 seconds (outside of the ED and ICU). 44% of patients had hypertension, 17% had history of MI, and 7% had other evidence of CVD with stroke or heart failure. A Modified Early Warning Score (MEWS), an indicator for severity of illness, was documented on just 77% of patients but in those documented, 68% had the highest severity MEWS four hours prior to arrest, indicating a poor prognosis.

Otieno et al demonstrated a mortality rate of 30% (n=14/47) in DKA patients presenting to KNH in a prospective, descriptive analysis conducted between 2001-2002 (14). Primary determinants of mortality were abnormal renal function which was present in 71% of those that died, altered mental status (100%), a new DM diagnosis (64%) and being female (64%). There was one cross-sectional study on diabetic retinopathy by Nkumbe et al(15) that had 71 subjects, which was conducted in a diabetes clinic in the largest referral hospital in Kenya, Kenyatta National referral hospital. They included all patients accessing care at the diabetes clinic with a recent diagnosis of 1 year or less, and used screening with color fundus photographs. 30.4 % of all the participants were found to have diabetic retinopathy while 8.2% had vision threatening retinopathy.

A case series on all cancer patients presenting to Tenwek hospital, a well-established mission hospital located in Western Kenya with a catchment of 400,000 patients, demonstrated esophageal cancer to be the most common cancer diagnosis (17). In addition, they found a disproportionately high prevalence of the disease in patients aged 30y or younger. A subsequent case series from the same hospital on “young” esophageal cancer patients, defined as 30y or younger, demonstrated that established risk factors for disease such as tobacco and alcohol are unlikely or absent in this population (16). Among patients diagnosed, no women (n=44) reported tobacco use, and only one reported alcohol use. Family history, on the other hand, was prevalent in 79% (n=45/60) of cases, with 44% reporting a first degree relative, and 28% with multiple relatives affected. The final study addressing cancer was also the only qualitative study included, and the only study that addressed attitudes towards care or care received. The study addressed ethical issues related to compulsory and voluntary screening in outpatient clinics in Kisumu (19).

They found that patients appreciated education associated with pre-testing, desired immediate results rather than waiting, demonstrated anxiety over results and coping with positive screening results, and desired verbal over written results given anxiety associated with the impersonal nature of written results, and illiteracy.

Finally, we identified a sole study on renal disease conducted by Wools-Kaloustian et al(18) which assessed antiretroviral naïve HIV-infected outpatient population in Western Kenya. It was a cross sectional study with 373 participants. Of the 373 participants, 43 (11.5%) were found to have renal insufficiency with a creatinine clearance of <50(18).

## DISCUSSION

In this study, we found a dearth of literature on NCDs in the acute and emergency care setting in Kenya. Of all of the studies, we found that the vast majority used a quantitative study design, and around half conducted retrospective chart reviews as part of their methodology. Of the leading four NCDs(3), chronic respiratory disease was unaddressed. Two studies were helpful in providing initial capacity assessments, though they were limited in their location and scope (10, 11). Another two studies addressed cardiovascular disease morbidity and mortality (12, 13), the foremost contributor to the global NCD burden, albeit in a tertiary care center that is private and unlikely to be accessed by majority of the population. None of the studies addressed interventions, treatment or implementation science targeting acute or emergency care populations in Kenya. Finally, all of the studies were observational in nature, no clinical trials were found.

Notably, five out of eight of the facility-based studies were conducted on patient populations at Kenyatta National Hospital, and Moi Teaching and referral hospital (12-15, 18). As two of the major referral hospitals in the country, both have access to large patient populations, and academic physicians with the potential resources to facilitate conducting research. Reasons for lack of research from other sites may also be due to publication bias(20).All the same, the lack of studies from other facility levels, and regions, particularly in rural regions where the majority of care is provided (7) needs to be addressed. In particular, nationally representative studies are needed.

In addition, there is a need for long-term data collection through the use of cohort studies, surveillance and registries (21) to assess epidemiology, mediators of disease, and outcomes of patients to better equip the healthcare system for resource allocation based on trends. This is particularly true for those diseases with a large burden of morbidity (3), such as cardiovascular disease, asthma and chronic kidney disease.

Majority of the study designs that met our inclusion criteria used quantitative methods, and we found only one qualitative study. Given the predominance of retrospective chart reviews in clinical research originating from LMICs (22)due to a lack of access to data, and other barriers that hinder de novo collection of primary data, this is not surprising. All the same, prioritization should also be given to conducting mixed methods or qualitative studies, which are lacking. The value for qualitative data tends to commonly be overlooked(23, 24). Qualitative studies are needed to better understand the underlying contributors to patient and provider practices pertaining to NCDs in the acute and emergency care setting. Implementation science will also be important in determining approaches to intervention development, piloting and dissemination (25). Finally, randomized controlled trials assessing effectiveness of clinical therapy, guidelines and other interventions are desperately needed in emergency care populations to guide best-practices unique to this setting (26).

In addition, most of the studies focused on patient clinical outcomes, with only three assessing healthcare system factors, such as resource availability (10, 11)and provider compliance (12). In the study by Wachira et al on STEMI patients at AKUHN (12), they found that compliance with guidelines in early phases of CVD care, such as obtaining EKGs on patients, was high likely secondary to the recent implementation of guidelines prior to the study (12). The authors cite the relatively new implementation of PCI at their institution, and difficulty accessing PCI outside of regular business hours, as limitations to compliance with definite therapy (PCI). In the second study by Wachira et al, which assessed IHCA patients (13), they found that a quarter of patients that arrested did not have a MEWS score documented on the chart. They purport that enforcing documentation, and indicators such as the MEWS to help identify high risk patients early on may help mobilize resources earlier for patients that need them and in turn reduce morbidity and mortality from delayed recognition (13). Implementation of healthcare system interventions has been shown to improve outcomes, such as with pre-hospital guidelines for CVD outcomes (27), early blood pressure control in hemorrhagic strokes (28), specialty training for pediatric airway management (29), and diabetes referral to care guidelines (30) in both Western nations with well-established emergency care, such as the US and Europe, and other African settings. Greater attention is needed addressing healthcare system interventions, clinical policy development, guideline implementation, and assessment of evidence-based, locally-generated quality measures(31, 32).

While it is encouraging that studies focused on the leading NCDs (3), CVD, cancer, and diabetes, no studies addressed chronic lung disease. Furthermore, we found only one study on complications of disease, which was assessing diabetic retinopathy (15). In line with the WHO Global NCD action plan (3), we believe that given the morbidity and mortality of these leading four chronic NCDs, studies should be aligned with these diseases. The research gap on chronic lung conditions has been identified as a key priority area with need for advanced research in Africa, including due to the unique exposure in LMICs associated with biomass fuel (33, 34). Furthermore, asthma in pediatric patients is a significant source of morbidity (35). More research is also needed on NCD-related complications such as stroke, diabetic retinopathy, kidney failure, and diabetic neuropathy, to further guide resource allocation and interventions targeting these patients when they present to the ED (15, 36-38). Moreover, the studies by Dawsey et al (16)and White et al (17) reinforce the need for primary data in the LMIC setting. As demonstrated, factors such as hot tea and diet have been shown to be leading risk factors associated with esophageal cancer in Kenyan populations, a deviation from more commonly established factors like tobacco and alcohol(3). In sum, greater understanding of the burden of disease, contributing factors, and complications of disease is desperately needed to help mitigate mortality and morbidity (39, 40).

## LIMITATIONS

Emergency medicine is a fledgling specialty in most parts of the world including Africa (6) where, for example in Kenya, there has only been a single emergency medicine “consultant” (“attending”) in practice over the last decade. Lack of awareness on the relevance of emergency care is likely a major contributor to the lack of prioritization and, in turn, dearth of research on this population. In our own findings, the majority of ED-based literature has been spearheaded by the sole emergency medicine “attending” or “consultant” in the country (7, 11-13). This demonstrates that by prioritizing the training of emergency care professionals, we not only improve clinical outcomes for patients in the emergency care setting (6, 39), but in turn secondarily contribute to driving research and development of evidence-based guidelines for this population. Given increased visibility on the development of emergency care in recent years, such as via incorporation into the Universal Health Coverage agenda by the WHO(40), and efforts by the African Federation for Emergency Medicine and other such bodies (41), ideally research efforts will only increase in volume and quality.

Furthermore, limitations due to publication bias likely also contributed to the dearth of studies found in our study. Publications not indexed in databases from which we sourced our review may have been excluded. We attempted to mitigate these effects by incorporating grey literature, and reviewing Africa-centered databases such as Africa-Wide Information in the protocol. Additionally, we included all articles in the final thesis as long as they met our a priori inclusion criteria, on epidemiology, interventions and management (9).

Finally, data in the studies discussed are not representative of the general Kenyan population. Nearly half incorporate data from private hospitals and one mission hospital (n=4/10), which are not accessed by the majority of Kenyans, or assessed urban populations. The majority of Kenya’s population resides in the rural areas. More research needs to be done focusing on emergency care at lower acuity health facilities and in rural areas, as recommendations made based on findings among urban populations cannot necessarily be generalized nationally.

## CONCLUSION

There is limited evidence on epidemiology, interventions and management of NCDs in Kenyan acute and emergency care populations, and there is need for increased research. Opportunities for research include nationally representative epidemiological studies on NCDs, and associated complications, as outlined by the WHO NCD action plan with the consideration for long-term cohort studies, registries and surveillance. Furthermore, studies are needed to address health system factors affecting NCD care in these settings, including through use of implementation science to inform policy, guideline development and dissemination. Finally, randomized, controlled trials to assess current and novel interventions among these populations are needed widely in LMIC emergency care settings, including Kenya.

## Data Availability

The authors confirm that the data supporting the findings of this study are available within the article and its supplementary materials.

